# Analysis of immunization, adverse events, and efficacy of a fourth dose of BNT162b2 vaccine

**DOI:** 10.1101/2022.04.05.22273434

**Authors:** Maria Elena Romero-Ibarguengoitia, Arnulfo González-Cantú, Diego Rivera-Salinas, Yodira Guadalupe Hernández-Ruíz, Ana Gabriela Armendariz-Vázquez, Irene Antonieta Barco-Flores, Rosalinda González-Facio, Miguel Ángel Sanz-Sánchez

**Affiliations:** Research Department, Hospital Clínica Nova de Monterrey; Avenida del Bosque 139, Cuauhtémoc, Cuauhtémoc, 66450, San Nicolás de los Garza, Nuevo León, México; Vicerrectoría de Ciencias de la Salud, Escuela de Medicina, Universidad de Monterrey, Avenida Morones Prieto, 4500-Pte, Zona Valle Poniente, 66238, San Pedro Garza García, Nuevo León, México

**Keywords:** Coronavirus, COVID-19, vaccination, immunization, adverse effects, antibodies, fourth dose

## Abstract

**Importance:** Scarce information exists concerning the seroconversion and adverse events after immunization (AEFI) of the fourth dose of a SARS-COV-2 vaccine.

**Objective:** Correlate the magnitude of the antibody response to vaccination with previous clinical conditions and AEFI of the fourth dose of BNT162b2 mRNA.

**Design:** Observational study where SARS-CoV-2 spike 1-2 IgG antibodies IgG titers were measured 21-28 days after the exposition of the first, and second dose, three months after the second dose, 1-7 days after the third dose, before the fourth dose, and 21-28 days after the fourth dose of BNT162b2 mRNA.

**Setting:** The study was conducted on healthcare workers of a private hospital in Northern Mexico.

**Participants:** Inclusion criteria were healthcare workers of both genders, any age, who planned to conclude the immunization regimen. The exclusion criteria were previously given any SARS-CoV-2 vaccine prior to study entry.

**Intervention:** Subjects were exposed to four doses of the BNT162b2 mRNA vaccine.

Main Outcome and Measures:

The anti-S1 and anti-S2 IgG antibodies against SARS-CoV-2 in plasma samples were measured with chemiluminescence immunoassay developed by DiaSorin.

**Results:** We recruited 112 subjects [43 (SD 9) years old, 74% women].

After the first dose, subjects had a median (IQR) AU/ml IgG of 122(1904), with an increase to 1875 (2095) after the second dose, 3020 (2330) after the third dose, and 4230 (3393) after 21-28 of a fourth dose (p<0.01). The number (%) of any AEFI between doses was 90 (80.4), 89(79), 65(58), 69 (61.5), after first, second, third, and fourth, respectively, p<0.001. After the fourth dose, the most frequent AEFI was pain at the injection site (87%). Fever was slightly more frequent after the third and fourth doses, 9 (13.8) and 8 (11.4%) cases, respectively, and adenopathy was more frequent after the fourth dose [in11(15.7%) cases]. There was a correlation between AEFI in the fourth dose with gender and antibody levels (p<0.05). The highest proportion of AEFI was considered mild after the fourth dose. During the Omicron outbreak, 6 (5.3%) had mild SARS-CoV-2 during 8-28 days of the fourth dose.

**Conclusions and Relevance:** The fourth dose of BNT162b2mRNA increases S1/S2 IgG 33.6 times with mild adverse events.

**Registration number:** NCT05228912

**Key points:** *Question:* What is the magnitude of antibody response to vaccination and adverse events after immunization (AEFI) of a fourth dose of BNT162b2 mRNA?

*Findings:* This cohort included 112 healthcare workers. We measured S1/S2 IgG vs. SARS-CoV-2 after the first, second, third and fourth dose. Compared to the first dose, antibodies increased 33.6 times the antibody levels after the fourth dose. We found minimal to moderate adverse events. The change in antibodies correlated with AEFI. During the Omicron outbreak 6 (5.3%) had mild SARS-CoV-2.

*Meaning:* A fourth dose of BNT162b2mRNA increases S1/S2 IgG with mild to moderate adverse events.

## Introduction

The SARS-CoV-2 worldwide pandemic has been a burden to any health system due to the agent’s high contagiousness and unpredictable outcome. Until March 2022 globally, SARS-CoV-2 has produced a total of 470.839,745 confirmed cases and 6,092,933 reported deaths according to the World Health Organization (WHO).^1^ This agent has proven to produce multiple waves of disease with more transmissible variants leading to long-term sequelae and perpetuating its exceeding impact on the economy and healthcare system.^2^

Given these circumstances, the development of vaccines against SARS-CoV-2 began urgently to mitigate its impact. In early 2021 multiple vaccines were created in order to prevent the infection of the disease.^3^ PfizerBioNTech (Pfizer Inc, New York, NY, USA, and BioNTech SE, Mainz, Germany) was one of the first laboratories to develop a mRNA vaccine against SARS-CoV-2, called BNT162b2.^4,5^

On the contrary, an unknown subject of interest is the duration of immunity generated by vaccination. Previous studies have shown a decline in antibody titers three to six months after the complete vaccination regimen. Khoury, et al. stated in their study a drop-out in antibody titers in four months following the complete vaccination regimen with only 6.3% of the maximal titer.^6^

As for today, The Centers for Disease Control and Prevention (CDC) has approved the application of a booster shot with BNT162b2 or mRNA-1273 (Moderna TX, Inc., USA; Rovi Pharma Industrial Services S.A., Spain) at least 5 months after the original vaccination scheme to everyone 12 and 18 years and older, respectively.^7^ Furthermore, for immunocompromised patients, CDC recommends a three-dose vaccination scheme with a booster 3 months after the third dose.^8^

At the beginning of 2022, Pfizer and BioNTech applied to the Food and Drug Administration (FDA) for the approval of a fourth dose of BNT162b2 in order to enhance the vaccine’s efficacy in reducing the rates of infection and severe illness, especially in adults 65 years and older.^9^ The application of a fourth BNT162b2 dose in non-immunocompromised patients is less studied. Therefore, our study aimed to correlate the magnitude of the antibody response to vaccination with previous clinical conditions and adverse events following immunization (AEFI) of a fourth dose of BNT162b2 mRNA in health workers. ^10,11^ Our secondary aim was to describe the incidence of SARS-COV-2 infection.

## Materials and methods

This study was a prospective observational cohort that followed healthcare workers who received a complete vaccination scheme with BNT162b2 vaccine in 2021, plus a third and fourth BNT162b2 booster 4 to 6 months after each last dose, respectively. This study was conducted in a Hospital in Northern Mexico. The study followed STROBE guidelines.^12^ Our study received approval from the local Institutional Review Board /Ref.;26022021-CN-1e-CI) and was conducted per the Code of Ethics of the World Medical Association (Declaration of Helsinki) for experiments that involve humans.

The inclusion criteria were healthcare professionals of both genders and any age who consented to participate, with a complete BNT162b2 national vaccination regimen, the application of the third BNT162b2 dose 4-6 months after the original vaccination scheme, and an additional BNT162b2 booster 4-6 months following the third dose. Exclusion criteria were subjects vaccinated prior to the study entry, heterologous vaccination regimen, and the application of any booster with any COVID-19 vaccine but BNT162b2.

Initially, the participants were recruited when the Health System started the vaccination campaigns. We invited the healthcare workers who intended to complete the BNT162b2 scheme to participate in the study. Inclusion-exclusion criteria were applied. Every participant signed an informed consent where the study was explained. A plasma blood sample was collected for every follow-up, and a questionnaire was applied. The concern about SARS-COV-2 variants (Delta and Omicron) led participants to inform us of applying a third and fourth BNT162b2 booster obtained by their means. Since our research protocol was approved after participants received the first dose, the blood samples were taken 21-28 days after the first and second BNT162b2 dose (T1 and T2, respectively), 3 months after the second dose (T3), 1-7 days after the third dose (T4), 21-28 days following the third BNT162b2 dose (T5), 3 months after the third dose (T6), prior the fourth dose’s application (T7), and 21-28 days following the fourth BNT162b2 dose (T8).

The questionnaires applied in every follow-up aimed to recover information about the patient throughout the whole study. The first questionnaire recovered the patient’s medical history and previous SARS-CoV-2 infection. The information recovered in the questionnaires from the 21-28 days after applying the four BNT162b2 doses follow-up included AEFI and SARS-CoV-2 infections after each dose.

### Primary outcome and IgG determination

Our main outcome was observing the humoral response elicited by vaccination through the measurement of specific antibodies against SARS-CoV-2 and correlating the results with previous SARS-CoV-2 infection, AEFI, and clinical conditions. Specific anti S1-S2 IgG antibodies against SARS-CoV-2 were measured by laboratory personnel using a chemiluminescence immunoassay (CLIA) by DiaSorin. This assay had a sensitivity of 97.4% (95% CI, 86.8-99.5) and a specificity of 98.5% (95% CI, 97.5-99.2). The results were interpreted as follows: < 12.0 AU/ml as a negative result; 12.0 to 15.0 AU/ml as indeterminate; and > 15.0 AU/ml as a positive result.

The variables we analyzed were gender, age, previous medical history (e.g., hypertension, type 2 diabetes mellitus, asthma, obesity, obstructive pulmonary disease, any heart condition, liver conditions, cancer, smoking, autoimmune diseases), previous SARS-CoV-2 confirmed infections (through nasal swab or serologic tests). The AEs reported with any BNT162b2 dose (i.e., pain at the injection site, fever [> 37.5ºC], headache, adenopathy).

### Statistical Methods

The sample size included all the healthcare workers that self-reported receiving a third and fourth dose of BNT162b2. The researchers reviewed the quality control and anonymization of the database. Normality assumption was evaluated with the Shapiro-Wilk test or Kolmogorov. Log10 transformations were used when it was appropriate. We used descriptive statistics for the analysis, such as frequencies, percentages, median, interquartile ranges, mean, and standard deviation. We performed a Friedman test to compare the anti S1 and S2 antibody titers through time and a Mann Whitney U test to compare antibody levels between subjects previously exposed to SARS-CoV-2. Cochran’s Q test was used to compare AEFI between vaccine doses.

We developed a mixed model where the dependent variable was the antibody titers. The reference group was the anti S1-S2 IgG antibody titers 21-28 days following the first BNT162b2 dose. We performed an individual variation as a random effect, and the fixed effects were the time of every anti S1-S2 antibody, gender, and age. Finally, we computed Poisson generalized linear models where the counts of any AEFI after the fourth dose were the outcome variables. Included regressors were genders, SARS-CoV-2 history, and every time the antibody was measured (see method section).

Completely missing at random values (< 30%) were imputed using multiple imputations through regression models. SPSS version 25 and R v 4.0.4 was the statistical program used to analyze data. The analysis was two-tailed, and a p-value < 0.05 was considered statistically significant.

## Results

Twelve (112) recruited subjects were vaccinated with a fourth BNT162b2 shot, of which 83 (74%) were women. The group’s mean (SD) age was 43 (9) years. Most of the participants were nurses (n=64, 57%) and medical providers (n=20, 17%). The most common comorbidity reported was obesity (n=32, 28.6%), followed by hypertension (n=12, 10.7%), dyslipidemia (n=9, 8.0%), prediabetes (n=8, 7.1%), hypothyroidism (n=7, 6.3%), type 2 diabetes mellitus (n=6, 5.4%), and non-alcoholic fatty liver disease (n=4, 3.6%). Seven subjects (6.3%) stated active smoking, and 2 (1.8%) stated the use of immunosuppressive therapy. Supplementary Table 1 shows the medical history reported by the participants.

**Table 1.** Anti S1-S2 IgG antibody titers follow-up according to SARS-CoV-2 infection history

| **Time points** | **Total**<br>**(n=112)**<br>**AU/ml** | **Fold**<br>**increase<sup>1</sup>**<br>**(n=112)** | **Negative**<br>**SARS-CoV-2**<br>**infection**<br>**history (n=63)**<br>**AU/ml** | **Positive**<br>**SARS-CoV-2**<br>**infection**<br>**history (n=49)**<br>**AU/ml** | **p-value** |
| --- | --- | --- | --- | --- | --- |
| 21-28 days after first dose (T1) | 122<br>(1904.5) |  | 97 (71.0) | 2220 (6122.5) | < 0.001 |
| 21-28 days after second dose (T2) | 1875<br>(2095.0) | 14.36 | 1470 (1476.0) | 2690 (3950.0) | < 0.001 |
| Three months after second dose (T3) | 300 (544.9) | 1.45 | 216 (241.2) | 486 (1584.7) | < 0.001 |
| 1-7 days after third dose (T4) | 489<br>(1055.8) | 3.00 | 405 (1002.0) | 814 (1251.5) | 0.043 |
| 21-28 days after third dose (T5) | 3020<br>(2330.0) | 23.75 | 2840 (1810.0) | 3240 (2490.0) | 0.099 |
| Three months after third dose (T6) | 1035<br>(920.0) | 7.48 | 872 (825.0) | 1320 (1142.0) | 0.004 |
| Prior the application of fourth dose (T7) | 598 (728.5) | 3.90 | 468 (557.0) | 673 (1119.7) | 0.021 |
| 21-28 days after fourth dose (T8) | 4230<br>(3393.0) | 33.67 | 4150 (3560.0) | 4370 (3230.0) | 0.837 |
| p-value | < 0.001 | < 0.001 | < 0.001 | < 0.001 |  |

Data are presented in median and interquartile ranges. Friedman test was performed for comparison between timepoints. Mann Whitney U test was performed for comparisons between positive and negative SARS-CoV-2 history. A p<0.05 was considered statistically significant.
<sup>1</sup>The fold increase in the follow-up was compared to the antibody titers taken 21-28 days after the first dose.

### Antibody titers

The anti S1-S2 IgG antibody titers against SARS-CoV-2 showed significant changes in applying BNT162b2 doses and time. Median (IQR) antibody titers AU/ml after first dose was 122 (1904.5); 21-28 days after second dose, 1875 (2095.0); three months after two doses, 300 (544.9); 1-7 days after third dose, 489 (1055.8); 21-28 days after third dose, 3020 (2330.0); three months after third dose, 1035 (920.0); prior the fourth dose, 598 (728.5); and 21-28 days after the fourth dose 4230 (3393.0).

While comparing the antibody titers after 21-28 days after each dose against the ones reported after the first dose, there was a 14.36, 23.75, and 33.67-fold increase after the second, third, and fourth BNT162b2 doses, respectively (p<0.001). There was a seven-fold increase in antibody levels when compared 21-28 days after the fourth dose with 155(22) after the third dose levels (p<0.001). Finally, there was a statistical difference between antibody levels 21-28 days after the third and fourth dose (p<0.001).

On the other hand, three months after the second and third dose, there was a decrease of 84% and 65.7%, respectively. Before the fourth dose, 155 (22) days after the third dose, there was a decrease of 80.2%.

Subjects were divided on their SARS-CoV-2 history for antibody comparisons (n=63 and n=49, respectively). There were significantly higher results in the antibody titers with a positive SARS-CoV-2 infection history (p < 0.05), except for the titers from the 21-28 days after the third and fourth dose (p=0.099 and p=0.837, respectively). See Table 1 for antibody comparisons through time and previous SARS-CoV2. History. Figure 1 represents the anti S1-S2 IgG antibody titers in log 10 in the follow-up.

**Figure 1.**
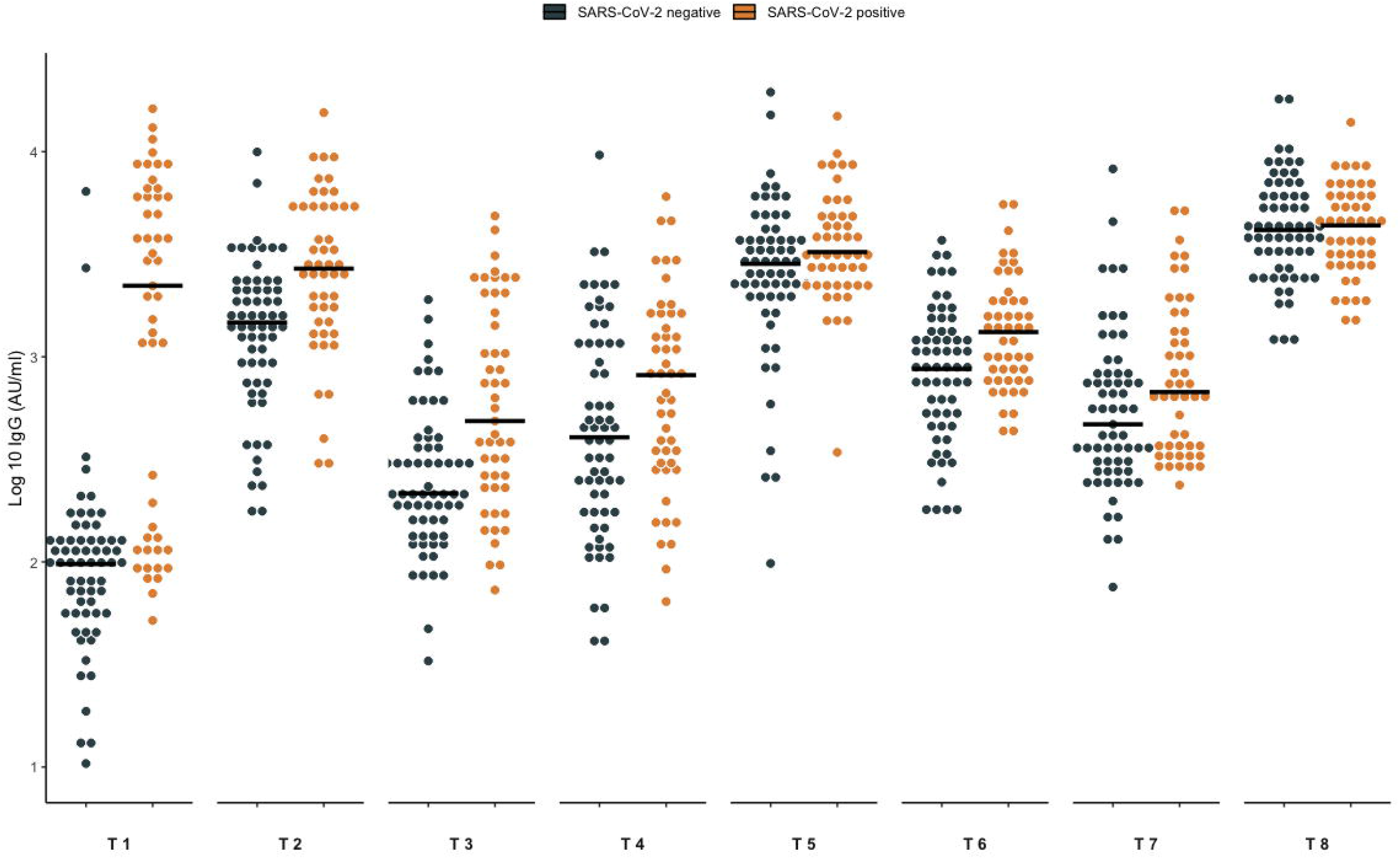
Anti S1-S2 IgG antibody titers according to SARS-CoV-2 infection history. The anti S1-S2 antibody titers are expressed in 10Log logarithm. The subjects were divided into positive and negative SARS-CoV-2 infection history. T1: 21-28 days after the first BNT162b2 dose. T2: 21-28 days after the second dose. T3: 3 months after the second dose. T4: 1-7 days after the third dose. T5: 21-28 days after third dose, T6: 3 months after third dose. T7: prior fourth dose. T8: 21-28 days after the fourth dose.

We created a mixed model where the anti S1-S2 IgG antibody titers were the dependent variable. The reference group for comparison was the antibody titers 21-28 days after the first dose. The antibody titers were analyzed in log10. This model found a significant positive effect in subjects with a SARS-CoV-2 infection history (β=0.32, p < 0.001). There was a positive effect after the application of the second, third, and fourth BNT162b2 doses (β= 0.79, β=1.0, and β= 1.2, respectively) (p < 0.001), in which the highest effect was due to the administration of the fourth dose.

There was a decline in the antibody titers in the three-month follow-up after the second and third doses. However, the titers were maintained above the antibody levels after the first BNT162b2 dose. The model did not show an effect on age or sex. Table 2. shows the mixed model of the anti S1-S2 IgG antibody titers.

**Table 2.** Mixed model of the anti S1 and S2 antibody titers against SARS-CoV-2

| | Estimate ($\beta$) | Std. Error | 95% CI | p-value |
| --- | --- | --- | --- | --- |
| Intercept | 0.27 |  |  |  |
| Age | 0.00 | 0.003 | -0.01, 0.00 | 0.15 |
| Sex | -0.05 | 0.065 | -0.18, 0.07 | 0.4 |
| SARS-CoV-2 infection before vaccination | 0.32 | 0.057 | 0.21, 0.43 | < 0.001 |
| 21-28 days after second dose | 0.79 | 0.047 | 0.70, 0.88 | < 0.001 |
| Three months after second dose | 0.10 | 0.047 | 0.00, 0.19 | 0.044 |
| 1-7 days after third dose | 0.27 | 0.047 | 0.18, 0.36 | < 0.001 |
| 21-28 days after third dose | 1.0 | 0.047 | 0.91, 1.1 | < 0.001 |
| Three months after third dose | 0.55 | 0.047 | 0.46, 0.64 | < 0.001 |
| Prior fourth dose | 0.35 | 0.047 | 0.25, 0.44 | <0.001 |
| 21-28 days after fourth dose | 1.2 | 0.047 | 1.1, 1.3 | < 0.001 |

Mixed model where the dependent variable was the anti S1 and S2 antibody titers against SARS-CoV-2. Antibody titers are analyzed as log 10. Antibody titers after 21-28 days after the first BNT162b2 dose were the reference group. p-value < 0.05 was considered statistically significant.

### Adverse Events Following Immunization (AEFI)

We evaluated the AEFI reported after each four doses. Less AEFI were reported after the third and fourth doses (first n=90, 80.4%; second n=89, 79.0%; third n= 65, 58.0%; and fourth dose n=69 61.6%, respectively). The time of appearance of the AEFI after the first and second doses were predominantly stated in the first four hours after the shot (n=73, 81.1% and n=41, 46.1%, respectively) (p < 0.001), whereas after the third and fourth, AEFI appeared in 5 to 24 hours after vaccination (n=38, 58.5% and n=36, 51.4%, respectively) (p < 0.001).

The most common AEFI related to the four BNT162b2 shots was pain at injection site (n=80, 88.9%; n=80, 89.0%, n= 52, 80.0%; and n=61, 87.0%, respectively), followed by headache (n=34, 37.8%; n=38, 42.7%; n=26, 40.0%; and n=33, 47.1%, respectively), and tiredness (n=22, 19.6%; n=39, 43.8%; n=27, 41.5%; and n=29, 41.4%, respectively). When comparing AEFI after each dose, adenopathy was proportionally more frequent after the fourth dose (p < 0.001), and fever after the third (p=0.036), whereas arthralgias was proportionally lower in the first those, than after the subsequent shots (p=0.019).

The severity of the AEFI was categorized as very mild, mild, moderate, severe, and very severe. The severity in the two-dose scheme was reported as very mild (first 60, 78.9% and second dose 42, 47.7%) (p= 0.01). However, the severity was mild after the third and fourth shots (24, 37.5%, and 36, 51.4%, respectively). See Table 3

**Table 3.** Adverse events following immunization after each administered BNT162b2 dose

| **AEFI (n=112)** | **First dose (%)** | **Second dose (%)** | **Third dose (%)** | **Fourth dose (%)** | **p-value** |
| --- | --- | --- | --- | --- | --- |
| Presence of any AE | 90 (80.4) | 89 (79.0) | 65 (58.0) | 69 (61.6) | < 0.001 |
| **Time of appearance** |  |  |  |  |  |
| First four hours after | 73 (81.1) | 41 (46.1) | 8 (13.8) | 22 (31.4) | < 0.001 |
| Five to 24 hours after | 5 (4.5) | 24 (27.0) | 38 (58.5) | 36 (51.4) | < 0.001 |
| Two to three days after | 12 (13.3) | 22 (24.7) | 17 (26.2) | 12 (17.1) | < 0.001 |
| Four to 7 days after | 0 | 1 (1.1) | 1 (1.5) | 0 |  |
| Seven to 10 days after | 0 | 1 (1.1) | 0 | 0 |  |
| **Symptoms** |  |  |  |  |  |
| Pain at injection site | 80 (88.9) | 80 (89.0) | 52 (80.0) | 61 (87.0) | 0.838 |
| Adenopathy | 1 (1.1) | 1 (1.1) | 7 (10.8) | 11 (15.7) | < 0.001 |
| Fever (>38°C) | 4 (4.4) | 1 (1.1) | 9 (13.8) | 8 (11.4) | 0.036 |
| Arthralgias | 5 (5.6) | 25 (28.1) | 14 (21.5) | 16 (22.9) | 0.019 |
| Headache | 34 (37.8) | 38 (42.7) | 26 (40.0) | 33 (47.1) | 0.303 |
| Tiredness | 22 (19.6) | 39 (43.8) | 27 (41.5) | 29 (41.4) | 0.110 |
| Myalgias | 7 (7.8) | 26 (29.2) | 16 (24.6) | 15 (21.4) | 0.135 |
| Local edema or erythema | 5 (5.6) | 14 (15.7) | 6 (9.2) | 8 (11.4) | 0.432 |
| Feverish (37.5°-37.9°C) | 4 (4.4) | 10 (11.2) | 6 (9.2) | 12 (17.1) | 0.188 |
| Nausea | 4 (4.4) | 6 (6.7) | 3 (4.6) | 4 (5.7) | 0.392 |
| Palpitations | 3 (3.3) | 2 (2.2) | 3 (4.6) | 2 (2.9) | 0.836 |
| Nasal congestion | 3 (3.3) | 10 (11.2) | 3 (4.6) | 4 (10) | 0.095 |
| Diarrhea | 2 (2.2) | 4 (4.5) | 2 (3.1) | 1(1.4) | 0.392 |
| Pruritus | 2 (2.2) | 5 (5.6) | 1 (1.5) | 3 (4.3) | 0.682 |
| Ocular pain | 2 (2.2) | 0 | 0 | 0 | - |
| Insomnia | 2 (2.2) | 1 (1.1) | 2 (3.1) | 0 | 0.733 |
| Localized skin rash | 1 (0.9) | 2 (2.2) | 0 | 1 (1.4) | 0.572 |
| Vomiting | 1 (1.1) | 1 (1.1) | 0 | 2 (2.9) | 0.572 |
| Back pain | 1 (1.1) | 0 | 0 | 0 | - |
| Achromatopsia | 1 (1.1) | 0 | 0 | 0 | - |
| Chest pain | 0 | 4 (4.4) | 1 (1.5) | 2 (2.9) | 0.392 |
| Hypotension | 0 | 0 | 1 (1.5) | 1 (1.4) | - |
| Diffuse skin rash | 0 | 0 | 1 (1.5) | 0 | - |
| Systemic edema | 0 | 1 (1.1) | 0 | 0 | - |
| **Severity** |  |  |  |  |  |
| Very mild | 60 (78.9) | 42 (47.7) | 21 (32.8) | 20 (28.6) | 0.01 |
| Mild | 6 (7.9) | 27 (30.7) | 24 (37.5) | 36 (51.4) | 0.01 |
| Moderate | 10 (13.2) | 18 (20.5) | 17 (26.6) | 12 (17.1) | 0.01 |
| Severe | 0 | 1 (1.1) | 1 (1.6) | 2 (2.9) | - |
| Very severe | 0 | 0 | 1 (1.6) | 0 | - |

Data are presented in frequencies and percentages. Cochran's Q test was performed for comparison. A p-value < 0.05 was considered statistically significant. AEFI: adverse events following immunization.

We computed Poisson generalized linear models to evaluate the predictors related to the appearance of any AEFI after the fourth dose of BNT162b2. Women correlated with more counts of AEFI (Log IRR-0.59, p < 0.001). Also, there was a small effect on antibody levels (Log IRR. 0002). A marginal effect was on SARS-CoV-2 history (Log IRR 0.16, p= 0.059). See Table 4.

**Table 4.** Poisson generalized linear models of factors related to adverse events following immunization

| Variable | log(IRR) | 95% CI | p-value |
| --- | --- | --- | --- |
| Gender | -0.59 | -0.82, -0.37 | < 0.001 |
| SARS-CoV-2 infection before vaccination | 0.16 | -0.01, 0.33 | 0.059 |
| Antibody titers | 0.0000292 | 0.0000003.00005 | 0.048 |

IRR: Incidence Rate Ratio, CI: Confidence Interval. Poisson generalized linear regression was performed to analyze the factors related to adverse events following immunization. p-value < 0.05 was considered statistically significant.

### SARS-CoV-2 infection

We considered a positive SARS-CoV-2 infection history when a subject had a confirmatory swab or serologic test. Throughout the whole follow-up, 58 (51.8%) subjects reported at least one SARS-CoV-2 infection, of which 43 (38.4%) were infected just once, 14 (12.5%) two times, and 1 (0.9%) three times.

Before the vaccination scheme, when the original variant caused infections, 49 (43.8%) subjects stated an infection, of which 47 (95%) received ambulatory treatment, 2 (4%) were hospitalized, and 1 (2%) was admitted to an Intensive Care Unit (ICU). Between the first and second dose, 1 (0.9%) participant was infected through the first wave and original variant. Between the second and third dose appeared the alfa, beta, gamma, and epsilon variants, in which just 1 (0.9%) subject was infected. The third wave caused by the delta variant arrived between the third and fourth dose, producing 6 (5.4%) cases, all infected after eight days of receiving the vaccine and 2 of the five months after this dose. After the fourth dose in the omicron wave, 12 (10.7%) were infected; 6 (5.3%) were infected 8-28 days after the fourth dose. All SARS-CoV-2 infection cases reported after administering the first BNT162b2 received ambulatory treatment.

## Discussion

This study analyzed the effect of a fourth BNT162b2 dose regarding its immunogenicity by measuring anti S1-S2 IgG antibodies. Also, AEFI and SARS-CoV-2 infections were addressed. The major strengths of our study are the close follow-up of the population since they are health workers from the institution where the study was conducted.

The fourth dose showed a 33.6-fold increase compared to the titers after the first dose. There was a positive effect in antibody titers when subjects were previously exposed to SARS-COV-2. This effect faded when healthcare workers received the third and fourth doses. As previously stated, three to six months following the second BNT162b2 dose, the antibodies tend to decrease with time, and after three months of the third dose, there was the same effect.

Regev-Yochay et al. conducted a study where they found a 9-10 fold increase of antibody titers after a fourth dose but did not find any substantial difference between the third and fourth dose results. They concluded concluding that the fourth dose’s effect might peak the antibody titers rather than increase immunity^13^ In our study, we found a 33.6 fold increase in antibodies with the fourth dose compared to the first dose, and when comparing the increase with 4-6 months after the third dose, we found a seven-fold increase.

The difference in the increase of antibodies between studies could be related to the method used for antibody measurement. Also, we had higher mean antibody titers in the follow-up, which could have been due to the inclusion of patients with SARS-CoV-2 history. Finally, when comparing the antibody levels after the third and fourth dose, we found a statistical difference with an increase of 1.4-fold.

The fourth BNT162b2 shot was safe because there were no severe adverse effects of vaccination. Limited mild local and systemic AEFI were reported following the fourth dose with no major complications. After the four doses, the most commonly reported were pain at the injection site, headache, and tiredness. We found that women and the antibody titers correlated with the AEFI. On Regev-Yochay et al. reported no serious adverse events with no hospital admissions related to vaccination, concluding that the fourth BNT162b2 dose is safe as the previous shots.^13^ Although the most common AEFI in their study were fatigue, myalgia, and headache, they were low grade and limited AEFI as the ones reported in our study.

The previous study stated, reported that the cumulative incidence of SARS-CoV-2 infection with Omicron over the period from 8-29 days after the administered BNT162b2 shot was 18.3% compared to 25.3% (95% CI: 18.5 to 31.5%) among their control.^13^ Our study reported 10.7%, but 5.3% after the eight days following the fourth dose, which is a lower infection rate. Also, when comparing the infection rate after at least eight days after the third and fourth dose, there is a similar rate of SARS-CoV-2 infection, even though it is well known that the Omicron variant was more contagious and had a higher infection rate.^14,15^

Recently FDA and CDC have recommended a second booster in subjects older than 50 years or immunocompromised with at least four months of receiving the third boost.^16,17^ Other studies have suggested that a fourth dose has low efficacy in preventing mild or asymptomatic Omicron infections and that the infectious potential still exists there is an urgency of next-generation vaccine development. ^18,19^ We agree with previous publications; however, until a new generation of vaccines is available, a fourth dose regimen could be an option for elder and immunocompromised patients and younger and healthy subjects since our study showed it could prevent severe cases of SARS-CoV-2 and reduce hospital burden. subjects since our study showed it could prevent severe cases of SARS-CoV-2 and reduce hospital burden.

As for limitations of our study, this is an observational study, so we lack randomization and a control group; however, we were able to compare our information with previously published data. A larger sample size would be of interest for testing for other predictors of antibody change; however, the changes of antibody titers between doses were so different that we could achieve enough statistical power for the inferences addressed in the present manuscript. This study did not include basal antibody measurement because, unfortunately, we could not obtain these samples since our protocol was accepted after all participants had received the first dose. However, we think this manuscript is valuable since new SARS-COV2 variants are emerging, and scarce information exists concerning a fourth dose that could help prevent infection or severe cases. Future studies about new generation vaccines against SARS-CoV-2 are required to compare the results with this regimen.

In conclusion, the fourth dose of BNT162b2mRNA produced a humoral response by increasing 33.6 times S1/S2 IgG compared to one dose, with mild adverse events. Women had more AEFI after the fourth dose, and there is a small effect related to the antibody level. Even though the Omicron variant is contagious, we found a similar infection rate to the Delta variant after subjects received the fourth dose. All the infected cases were mild and treated ambulatory.

## Data Availability

All data produced in the present study are available upon reasonable request to the authors

## Acknowledgments

We thank the laboratory team, human resources, call center, technology, and maintenance personnel of Hospital Clinica Nova, for helping in the logistics of this research.

## Author Approval

all authors read an approved the final version of the manuscript

## Data Availability Statement

Data are available upon reasonable request to the authors.

## Ethics statement

Ethics committee/local Institutional Review Board from the school of Medicine from Universidad de Monterrey gave ethical approval: Ref.:26022021-CN-1e-CI. Funding

## References

1. World Health Organization. WHO Coronavirus (COVID-19) Dashboard. Accessed March 15, 2022. https://covid19.who.int

2. World Health Organization. Strategy to Achieve Global Covid-19 Vaccination by mid-2022. Accessed March 15, 2022. https://www.who.int/publications/m/item/strategy-to-achieve-global-covid-19-vaccination-by-mid-2022

3. Velavan TP, Meyer CG. The COVID□19 epidemic. Trop Med Int Health. 2020;25(3):278-280. doi:10.1111/tmi.13383

4. Pfizer Inc. Pfizer-BioNTech COVID-19 Vaccine | Pfizer. Accessed March 17, 2022. https://www.pfizer.com/products/product-detail/pfizer-biontech-covid-19-vaccine

5. Polack FP, Thomas SJ, Kitchin N, et al. Safety and Efficacy of the BNT162b2 mRNA Covid-19 Vaccine. N Engl J Med. 2020;383(27):2603–2615. doi:10.1056/NEJMoa2034577

6. Khoury J, Najjar-Debbiny R, Hanna A, et al. COVID-19 vaccine – Long term immune decline and breakthrough infections. Vaccine. 2021;39(48):6984–6989. doi:10.1016/j.vaccine.2021.10.038

7. CDC. COVID-19 Booster Shot. Centers for Disease Control and Prevention. Published April 1, 2022. Accessed March 17, 2022. https://www.cdc.gov/coronavirus/2019-ncov/vaccines/booster-shot.html

8. CDC.COVID-19 Vaccination. Centers for Disease Control and Prevention. Published February 11, 2020. Accessed March 17, 2022. https://www.cdc.gov/coronavirus/2019-ncov/vaccines/recommendations/immuno.html

9. Tanne JH. Covid-19: Pfizer asks US regulator to authorise fourth vaccine dose for over 65s. BMJ. 2022;376:o711. doi:10.1136/bmj.o711

10. Chen M, Yuan Y, Zhou Y, et al. Safety of SARS-CoV-2 vaccines: a systematic review and meta-analysis of randomized controlled trials. Infect Dis Poverty. 2021;10(1):94. doi:10.1186/s40249-021-00878-5

11. Fan YJ, Chan KH, Hung IFN. Safety and Efficacy of COVID-19 Vaccines: A Systematic Review and Meta-Analysis of Different Vaccines at Phase 3. Vaccines. 2021;9(9):989. doi:10.3390/vaccines9090989

12. Cuschieri S. The STROBE guidelines. Saudi J Anaesth. 2019;13(Suppl 1):S31–S34. doi:10.4103/sja.SJA_543_18

13. Regev-Yochay G, Gonen T, Gilboa M, et al. 4th Dose COVID mRNA Vaccines’ Immunogenicity &amp; Efficacy Against Omicron VOC. Published online February 15, 2022:2022.02.15.22270948. doi:10.1101/2022.02.15.22270948

14. CDC. Omicron Variant: What You Need to Know. Centers for Disease Control and Prevention. Published March 29, 2022. Accessed March 25, 2022. https://www.cdc.gov/coronavirus/2019-ncov/variants/omicron-variant.html

15. Chen J, Wang R, Gilby NB, Wei GW. Omicron Variant (B.1.1.529): Infectivity, Vaccine Breakthrough, and Antibody Resistance. J Chem Inf Model. 2022;62(2):412–422. doi:10.1021/acs.jcim.1c01451

16. U.S. Food & Drug Administration. Coronavirus (COVID-19) Update: FDA Authorizes Second Booster Dose of Two COVID-19 Vaccines for Older and Immunocompromised Individuals. FDA. Published March 29, 2022. Accessed April 1, 2022. https://www.fda.gov/news-events/press-announcements/coronavirus-covid-19-update-fda-authorizes-second-booster-dose-two-covid-19-vaccines-older-and

17. CDC. Coronavirus Disease 2019. Centers for Disease Control and Prevention. Published March 29, 2022. Accessed April 1, 2022. https://www.cdc.gov/media/releases/2022/s0328-covid-19-boosters.html

18. Gazit S, Saciuk Y, Perez G, Peretz A, Pitzer VE, Patalon T. Relative Effectiveness of Four Doses Compared to Three Dose of the BNT162b2 Vaccine in Israel. Public and Global Health; 2022. doi:10.1101/2022.03.24.22272835

19. Alejo JL, Mitchell J, Chiang TPY, et al. Antibody Response to a Fourth Dose of a SARS-CoV-2 Vaccine in Solid Organ Transplant Recipients: A Case Series. Transplantation. 2021;105(12):e280–e281. doi:10.1097/TP.0000000000003934

