## Supplemental information for "Analysis of immunization, adverse events, and efficacy of a fourth dose of BNT162b2 vaccine"

**Supplementary Table 1. Medical History**

| **Medical history (n=112)** | **Frequency (%)** |
| --- | --- |
| Obesity | 32 (28.6) |
| Hypertension | 12 (10.7) |
| Dyslipidemia | 9 (8.0) |
| Prediabetes | 8 (7.1) |
| Hypothyroidism | 7 (6.3) |
| Smoking | 7 (6.3) |
| Type 2 Diabetes Mellitus | 6 (5.4) |
| Non-alcoholic fatty liver disease | 4 (3.6) |
| Asthma | 3 (2.7) |
| Rheumatoid arthritis | 2 (1.8) |
| Use of immunosuppressive therapy | 2 (1.8) |
| Atrial fibrillation | 1 (0.9) |
| Heart failure | 1 (0.9) |
| Coronary heart disease | 1 (0.9) |
| Stroke | 1 (0.9) |
| Gout | 1 (0.9) |
| Pregnancy | 1 (0.9) |

Data are presented in frequencies and percentages.
